# Socioeconomic inequalities in prevalence and development of multimorbidity across adulthood: findings from the MRC 1946 National Survey of Health & Development

**DOI:** 10.1101/2020.08.14.20174037

**Authors:** Amal R. Khanolkar, Nishi Chaturvedi, Valerie Kuan, Daniel Davis, Alun Hughes, Marcus Richards, David Bann, Praveetha Patalay

## Abstract

**Objective:** To estimate multimorbidity trajectories and quantify socioeconomic inequalities based on both childhood and adulthood socioeconomic position in the risks and rates of adult multimorbidity accumulation.

**Design:** Prospective longitudinal national birth cohort study.

**Methods:** Participants from the 1946 National Survey of Health and Development (NSHD) who attended the age 36 assessment in 1982 and any one of the follow-up assessments at ages 43, 53, 63 & 69 (N=3,723, 51% males). Information on 18 health conditions was based on a combination of self-report, biomarkers, health records and prescribed medications. We estimated multimorbidity trajectories and delineated socioeconomic inequalities (based on childhood and adulthood social class and highest education) in multimorbidity at each age and in longitudinal trajectories.

**Results:** Multimorbidity increased with age from 0.7 conditions on average at 36 years to 3.7 at 69 years. Multimorbidity accumulation was non-linear, accelerating with age at the rate of 0.08 conditions/year (95% CI 0.07-0.09) at 36-43 years to 0.19 conditions/year (95% CI 0.18-0.20) at 63-69 years. At all ages, the most socio-economically disadvantaged had 1.2 to 1.4 times greater number of conditions on average compared to the most advantaged. The most socioeconomically disadvantaged by each socioeconomic indicator experienced an additional 0.39 conditions (childhood social class), 0.83 (adult social class) and 1.08 conditions (adult education) at age 69 years, independent of all other socioeconomic indicators. Adverse adulthood socioeconomic status was associated with more rapid accumulation of multimorbidity, resulting in 0.49 excess conditions in partly/unskilled compared to professional/intermediate individuals between 63-69 years. Disadvantaged childhood social class, independently of adult socio-economic position, was associated with accelerated multimorbidity trajectories from age 53 onwards.

**Conclusions:** Socioeconomically disadvantaged individuals have both earlier onset and more rapid accumulation of multimorbidity resulting in widening inequalities into old age, with independent contributions from both childhood and adulthood socio-economic position.

## Introduction

The management of long-term chronic health conditions poses a serious challenge for public healthcare systems worldwide.^1 2^ Conditions associated with unhealthy lifestyles, such as obesity, hypertension, diabetes, and coronary heart disease are increasingly common, while greater life expectancies means individuals live longer with chronic disease. These conditions rarely occur in isolation, hence multimorbidity – two or more chronic conditions in an individual – is a rising consequence.^3^ Multimorbidity is associated with poorer quality of life, decreased work productivity, increased use of ambulatory and in-patient healthcare, polypharmacy, increased functional limitations and mortality.^4-9^ Socioeconomic inequalities in health are well established and socioeconomic disadvantage is associated with both a greater burden of multimorbidity and earlier onset. ^2 10-21^

However most studies on the epidemiology of multimorbidity, and specifically those that have investigated socioeconomic inequalities, have largely been cross-sectional in design,^2 4 12 13 15 17 22^ precluding research on 1) longitudinal development of multimorbidity within the same individuals across adulthood, 2) longitudinal effects of early and later life socioeconomic position on prevalence and development of multimorbidity.

Independent effects of socioeconomic disadvantage in childhood on multimorbidity development across the lifecourse is not known. This is an important gap in knowledge, as socioeconomic disadvantage in childhood has lasting impacts on health, independently of adult socioeconomic circumstances.^23 24^ Excluding childhood socioeconomic circumstances might underestimate the extent of the impact of socioeconomic disadvantage across the lifecourse.

Further, no studies to-date have investigated multimorbidity development across the lifecourse, particularly in association with lifetime socioeconomic position, as this requires individual level population-based longitudinal data. Given ageing related processes and health deterioration, it is expected that the rate of multimorbidity accumulation will accelerate over the lifecourse. Any socioeconomic differentials in the rate of disorder accumulation are important to establish as they help uncover whether the inequalities observed at older ages are driven by earlier onset, faster accumulation or both.

This study examines multimorbidity development across adulthood and into older ages and investigates in detail socioeconomic inequalities in multimorbidity trajectories in a prospective nationally representative birth cohort study. We estimated multimorbidity development in the same individuals over time and, importantly, estimate the independent effects of childhood and adulthood socioeconomic circumstances on multimorbidity prevalence and rate of accumulation.

## Methods

### Study participants and setting

This study analysed data from participants in the MRC 1946 National Survey of Health and Development (NSHD).^25^ The NSHD includes a socially stratified and nationally representative sample of 5,362 participants born in one week in March 1946 in England, Scotland and Wales and recruited at birth. Extensive social, anthropometric, medical, lifestyle, and biological data were collected either during home visits or at clinical research centres in later years by professional interviewers and research nurses. More information on the study is available from the study’s website: https://www.nshd.mrc.ac.uk/.

The analysis sample for this prospective longitudinal study included participants who attended the age 36 assessment in 1982 and any one of the follow-up assessments at ages 43, 53, 63 & 69 (total N=3,723, including 1,843 women [49.5%]).

Attrition: Of the original 5,362 study participants, the study team was still in contact with 3,163 (59%) at age 63 years. By this age, 718 (13.4%) had died, 594 (11.1%) had previously withdrawn from the study, 567 (10.6%) lived abroad and 320 (5.9%) had been untraceable for more than ten years.

### Conditions

Data for 18 selected common chronic conditions on which consistent information was collected across the adulthood sweeps are the focus of this study^26^. These included diabetes, dyslipidaemia, hypertension, obesity, coronary heart disease, stroke, cancer, anaemia, respiratory-, kidney-, gastrointestinal-, skin-disorders, arthritis, Parkinson’s disease, epilepsy, depression & psychosis. Data sources, sweeps at which information is available and definitions for each of the 18 conditions are listed in Supplemental Table 1.

### Chronicity

We used the Delphi technique^27^ – a structured communication tool – to give an informed consensus on which conditions would be considered chronic (i.e. once a participant was diagnosed with a condition then he/she would be considered to have the condition throughout follow-up). The Delphi team consisted of ten clinicians who received the same form (Supplemental Table 2). Each clinician was requested to indicate their decision and provide a brief explanation supporting their decision.

### Multimorbidity

All 18 conditions were coded as binary variables indicating whether a participant had a condition or not at each sweep. The multimorbidity score in this paper is defined as the unweighted sum of the number of conditions (out of 18 conditions) accumulated by an individual^2 14^.

For those conditions considered chronic (like diabetes or hypertension), a participant was coded to have the condition throughout follow-up once diagnosed (Supplemental Table 1). For all participants, we calculated a multimorbidity score at each age which indicated the number of conditions accumulated over time (i.e. across the five age sweeps) and was the main outcome.

### Socioeconomic position (SEP)

Childhood SEP was based on the father’s occupational social class, reported at age 11. We used two measures of SEP in adulthood; educational level attained at age 33 and occupational social class ascertained at age 43. Occupational social class was coded into 4 categories: 1. Professional and intermediate (reference group in analysis), 2. Non-manual, 3. Manual, and 4. Partly skilled/unskilled. Educational levels were ascertained by the Burnham classification. The original 9 categories were collapsed into 3 groups for analysis: 1. None or vocational courses, 2. School leaving certificate (GCE ‘O’ and ‘A’ levels) and 3. Degree level and higher (university education, reference group).

## Statistical analysis

We used frequencies, percentages (prevalence of each condition at all age sweeps for the full study sample), cross tabulations (mean differences in multimorbidity score across the five age sweeps by socioeconomic indicators), and graphical display for descriptive analysis. To visualise prevalence and associations between conditions at each age, networks were constructed using the R package “igraph” (conditions represented by nodes with sizes proportional to prevalence). Partial correlations between each of the 18 conditions are represented as ‘edges’ in networks (networks only included those edges with partial correlations with absolute values >0.05 and p-values<0.05). The width of an edge was proportional to the strength of the partial correlation between two conditions (with positive partial correlations >0.05 represented by grey edges, and negative correlations<-0.05 shown in red).

Socioeconomic patterning in multimorbidity at each of the five age sweeps was analysed using multivariable linear regression. We first ran models to test individual associations between each of the three indicators and the multimorbidity score. The fourth model was mutually adjusted for all three socioeconomic indicators in order to assess each of their associations independent of other indicators. All models were adjusted for sex.

Longitudinal change in multimorbidity trajectories were analysed using linear spline mixed-effects modelling. This accounts for repeated measurements (or clustering) of multimorbidity scores within individuals over time and individual-level deviations from an overall average trajectory, with a non-linear specification of the average trajectory using linear splines^28^. Given the data structure – cohort members are of the same age at each time point – knots were assigned at ages 43, 53, 63 and 69 giving 4 splines corresponding to: ages 36 to 43, 43 to 53, 53 to 63 and 63 to 69. The model with a random intercept plus random slopes for the individual spline coefficients had a superior fit and was used in all subsequent modelling. This breaks-up the average change in multimorbidity score into separate slopes for each of the four spline variables representing an age band (each slope coefficient indicating rate of change in multimorbidity score per year in each period). We used linear spline mixed-effects modelling to first estimate the population-average multimorbidity trajectory for the full sample. We then ran four models, each one having a different socioeconomic exposure of interest plus sex; Model 1: childhood social class, Model 2: adulthood social class, Model 3: adulthood educational level and Model 4: mutual adjustment for all three socioeconomic indicators.

To examine whether inequalities change over time, we tested for interactions between childhood or adulthood social class and the individual spline variables. The model for adulthood social class included interactions with all 4 spline variables whereas the one for childhood social class included interactions with the ages 53-63 and 63-69 variables only. These models were mutually adjusted for other socioeconomic indicators. Interactions were significant and multimorbidity trajectories were plotted at the group level (childhood and adulthood social class separately) for visualisation based on model 4 above. Interactions between spline variables and education were not significant and are not shown.

Missing data across the five age sweeps was addressed using multiple imputation assuming missing at random (using 25 imputed datasets). Frequencies and distributions of non-imputed and imputed variables were similar. All analyses were conducted using Stata version 15 (StataCorp LP, College Station, Texas).

## Results

Table 1 shows the prevalence of the 18 conditions across the five ages. Prevalence of most conditions increased with age (for example obesity and hypertension increased from 7% and 6% at age 36 to 34% and 52% at age 69 respectively). The proportion with no conditions decreased progressively from 52% at age 36 to 7% at age 69 (Figure 1). Figure 2 displays results from network analysis with node size representing relative prevalence of a condition, and edges representing associations between conditions at each of the five ages. These networks illustrate both the relative increase in prevalence of disorders over time, and the greater interconnectivity between conditions with age. For example, prevalence of hypertension increases with age, *and* its association with dyslipidaemia gets stronger over time. In contrast, although more individuals have cancer over time, cancer is not independently associated with experiencing any of the other conditions at most ages (indicated by no edges between cancer and other conditions). Similarly, skin conditions are independent of most conditions over time, with the exception of respiratory conditions.

**Table 1.** Prevalence (%) of conditions and multimorbidity at each of the five age sweeps in 3,723 participants from the 1946 National Survey of Health and Development.

| Condition | Age (prevalence [95% CI]) |  |  |  |  |
| --- | --- | --- | --- | --- | --- |
|  | Age 36 | Age 43 | Age 53 | Age 63 | Age 69 |
| **Obesity** | 6.9<br>(6-7.7) | 12.8<br>(11.6-13.9) | 25<br>(23.2-26.6) | 32.2<br>(30-34.4) | 34.4<br>(32-36.7) |
| **Hypertension** | 6.3<br>(5.4-7.1) | 13.3<br>(12.1-14.5) | 32.4<br>(30.7-34.1) | 44.8<br>(42.9-46.6) | 52<br>(50-53.9) |
| **Dyslipidaemia** | - | - | 9.1<br>(7.9-10.2) | 32.2<br>(30-34.3) | 47.8<br>(45.4-50) |
| **Diabetes** | - | 1.5<br>(1-1.9) | 3.2<br>(2.5-3.8) | 10.7<br>(9.4-11.9) | 16.3<br>(14.8-17.7) |
| **CHD** | - | 3.8<br>(3.1-4.5) | 11.6<br>(10.2-12.9) | 20.2<br>(18.5-22) | 22.8<br>(20.9-24.5) |
| **Stroke** | - | - | 2.5<br>(1.4-3.6) | 6.4<br>(4.7-8.2) | 10.3<br>(8.6-11.9) |
| **Osteoarthritis** | - | - |  | 9.4<br>(7.8-10.9) | 27.5<br>(25.1-29.8) |
| **Rheumatoid Arthritis** | - | - | 1.3<br>(0.5-2.2) | 4.2<br>(2.5-5.9) | 9.7<br>(7.3-12) |
| **Anaemia** |  |  | 9<br>(7.8-10.2) | 6.8<br>(5.2-8.5) | 12.5<br>(10.4-14.6) |
| **Skin disorders** | 10.8<br>(9.7-11.8) | 18.6<br>(17.3-20) | 19.1<br>(17.6-20.5) | - | 16<br>(14.1-18) |
| **Respiratory disorders** | 16.1<br>(14.8-17.4) | 28.4<br>(26.8-30) | 17.5<br>(15.9-19) | 20.4<br>(18.7-22.1) | 25.6<br>(23.7-27.5) |
| **Gastrointestinal disorders** | 2.3<br>(1.6-2.8) | 8.9<br>(7.8-9.8) | 8<br>(6.7-9.1) | 21<br>(18.6-22.7) | 28.3<br>(25.1-21.5) |
| **Kidney disorders** | 4.9<br>(4.2-5.6) | 13.8<br>(12.6-14.9) | 13.3<br>(11.8-14.8) | - | 5<br>(2.8-7.2) |
| **Parkinson's disease** | - | - | - | 2.6<br>(1.2-4) | 5.5<br>(3.8-7.1) |
| **Cancer** | 0.6<br>(0.3-0.8) | 1.7<br>(1.3-2.1) | 5.6<br>(4.8-6.3) | 14.2<br>(13.1-15.3) | 19.5<br>(18.3-20.9) |
| **Epilepsy** | 1.5<br>(0.9-1.9) | 2.1<br>(1.4-2.7) | 4.2<br>(2.9-5.4) | 3<br>(2.1-4) | 8.3<br>(6.7-9.9) |
| **Depression** | 18.4<br>(16.9-19.7) | 16.4<br>(15.2-17.7) | 23<br>(21.3-24.7) | 23.4<br>(21.7-25.1) | 24.5<br>(22.7-26.9) |
| **Psychotic disorders** | 0.9<br>(0.5-1.4) | 2<br>(1.4-2.7) | 3.8<br>(2.6-5.1) | 6<br>(4.4-7.6) | 8<br>(6.2-9.8) |
| **Multimorbidity (mean)** | 0.7<br>(0.6-0.7) | 1.2<br>(1.1-1.3) | 1.9<br>(1.8-2.0) | 2.6<br>(2.5-2.7) | 3.7<br>(3.6-3.9) |

Note: Empty cells indicate data for that condition was not collected at the particular age.

**Figure 1.**
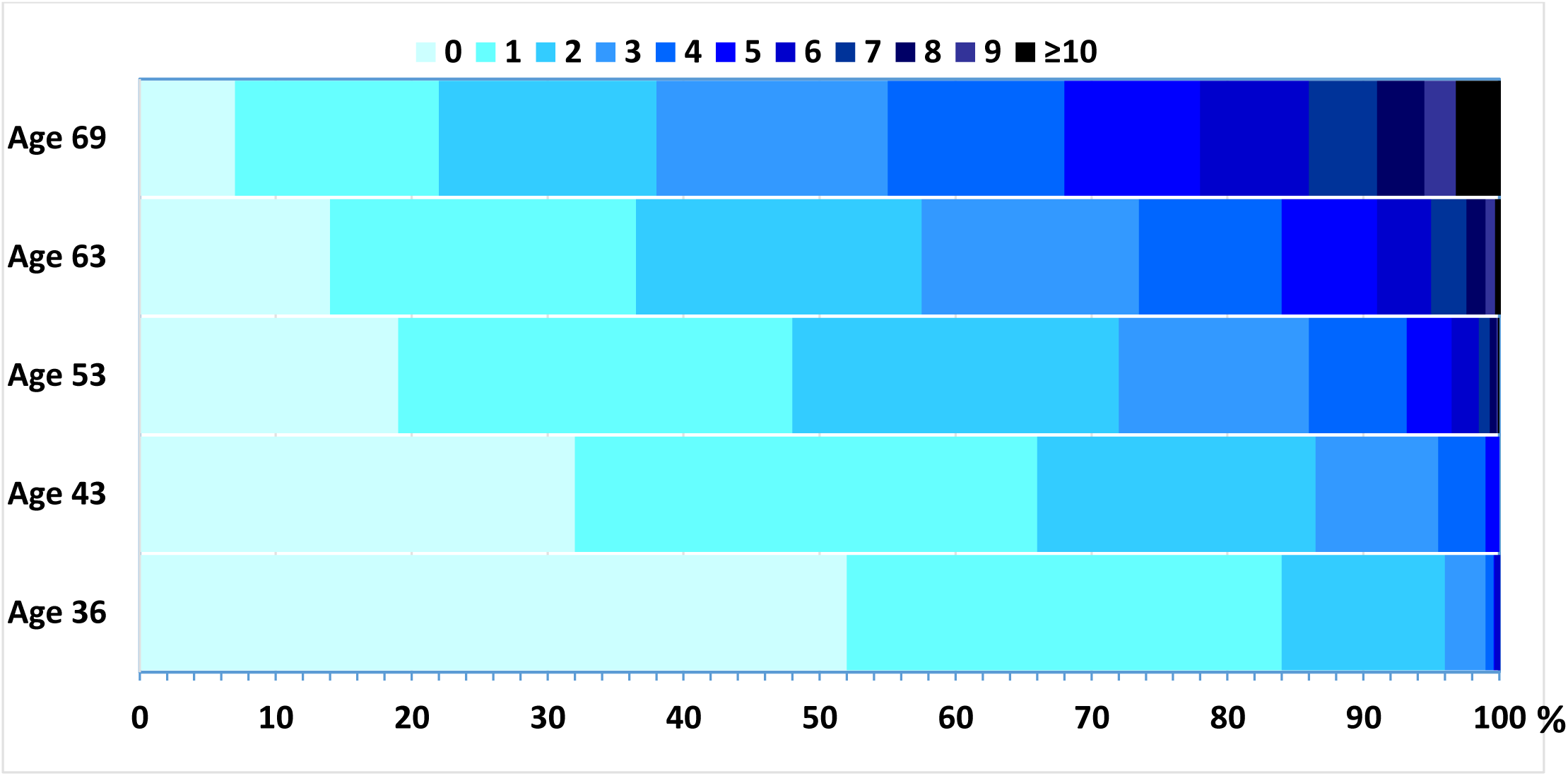
Percentage of participants with increasing levels of multimorbidity (based on 18 common conditions) at each age sweep in 3,723 participants from the 1946 National Survey of Health and Development.

**Figure 2.**
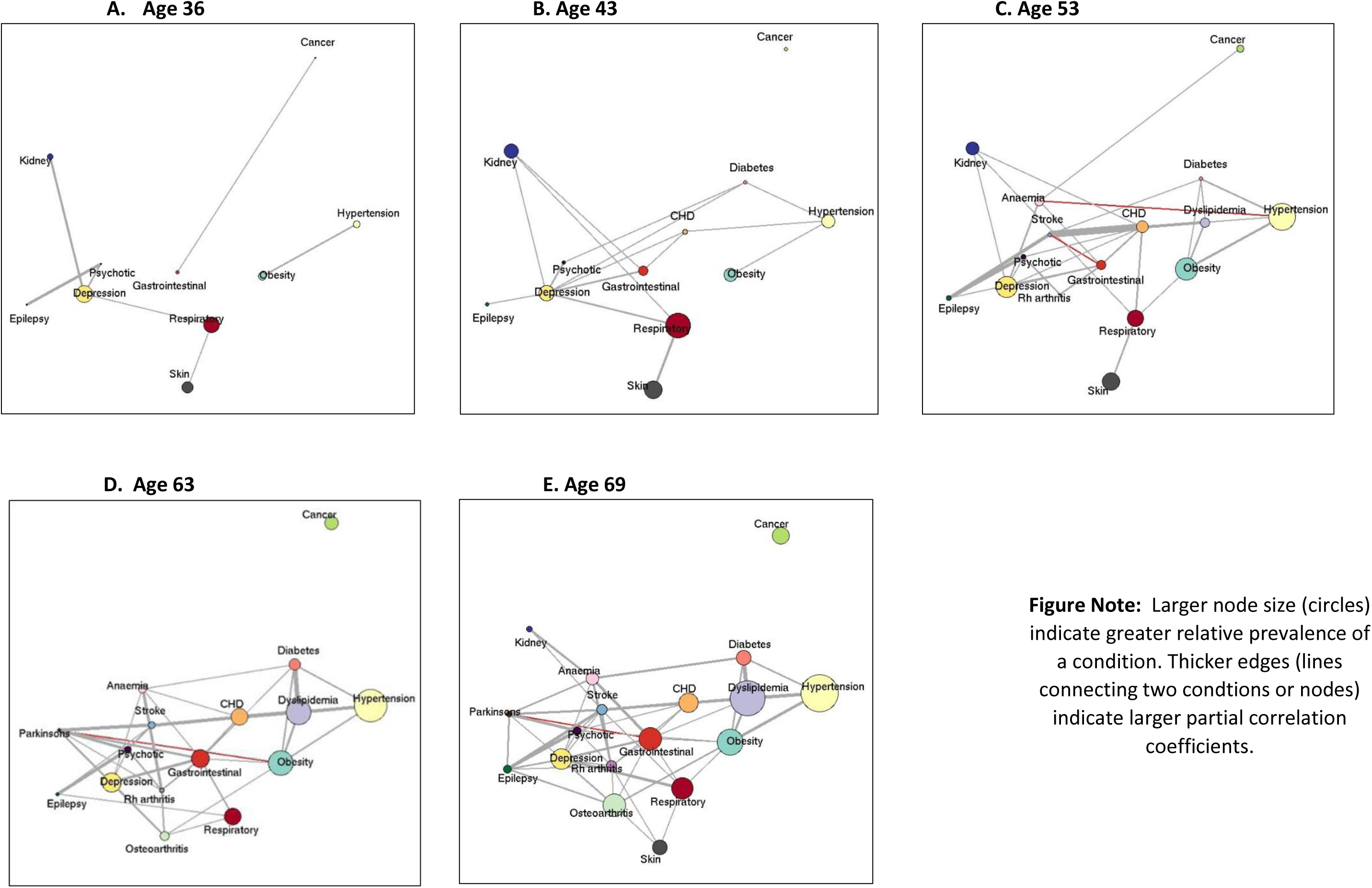
Network analysis illustrating relative prevalence (node size) of conditions and adjusted associations (edges) at each age in 3,723 participants from the 1946 National Survey of Health and Development. Larger node size (circles) indicate greater relative prevalence of a condition. Thicker edges (lines connecting two condtions or nodes) indicate larger partial correlation coefficients.

At each age sweep, socioeconomic disadvantage was associated with higher multimorbidity scores (Figure 3, Table 2 and Supplemental Figure 1, Supplemental Table 5). The mean difference in number of conditions between the most and least disadvantaged was observed with childhood and adulthood social class, and with educational level at all ages, and got progressively larger with age. At all ages, the most socioeconomically disadvantaged had a greater number of conditions on average compared to the most advantaged with the largest difference observed at age 69 with 1.4 more conditions (e.g. 0.60 vs. 0.85 at age 36, 1.67 vs. 2.28 at age 53 and 3.24 vs. 4.59 at age 69 for professional/intermediate vs. partly skilled/unskilled adulthood social class respectively, Supplemental Table 5). Compared with the most advantaged group (professional/intermediate), partly skilled/unskilled individuals had progressively higher multimorbidity scores across adulthood (Table 2, age 36:0.18, [0.08-0.28], age 69:0.83, [045-1.20]). Socioeconomic disadvantage in childhood and in adulthood were independently associated with higher levels of multimorbidity at ages 63 and 69 (Table 2, 0.39 [0.06-0.72] for partly skilled/unskilled childhood social class and 1.08 [0.67-1.49] for no education age 69). The most socioeconomically disadvantaged by each measure of SEP, adjusted for all other measures of SEP, experienced an additional 0.39 (childhood social class), 0.83 (adult social class) and 1.08 conditions (adult education) at age 69 years (Table 2).

**Table 2.** Results from linear regression modelling assessing associations between socioeconomic position and multimorbidity in 3,723 participants from the 1946 National Survey of Health and Development.

|  | Change in multimorbidity score<sup>a</sup> |  |  |  |  |  |  |  |  |  |
| --- | --- | --- | --- | --- | --- | --- | --- | --- | --- | --- |
|  | Age 36 |  | Age 43 |  | Age 53 |  | Age 63 |  | Age 69 |  |
| Covariates | β | 95% CI | β | 95% CI | β | 95% CI | β | 95% CI | β | 95% CI |
| **Childhood Social Class** |  |  |  |  |  |  |  |  |  |  |
| Professional/intermediate | Ref |  | Ref |  | Ref |  | Ref |  | Ref |  |
| Skilled non-manual | 0.03 | -0.07,0.12 | 0.08 | -0.05,0.21 | -0.01 | -0.18,0.17 | 0.11 | -0.13,0.35 | 0.18 | -0.13,0.50 |
| Skilled manual | 0.05 | -0.04,0.13 | 0.05 | -0.07,0.16 | 0.09 | -0.07,0.24 | **0.32** | **0.12,0.52** | **0.39** | **0.10,0.68** |
| Partly skilled/unskilled | 0.03 | -0.06,0.12 | 0.07 | -0.06,0.20 | 0.04 | -0.14,0.22 | **0.25** | **0.00,0.50** | **0.39** | **0.06,0.72** |
| **Adulthood Social Class** |  |  |  |  |  |  |  |  |  |  |
| Professional/intermediate | Ref |  | Ref |  | Ref |  | Ref |  | Ref |  |
| Skilled non-manual | 0.04 | -0.04,0.13 | 0.03 | -0.09,0.15 | 0.12 | -0.04,0.27 | 0.05 | -0.15,0.25 | 0.11 | -0.17,0.39 |
| Skilled manual | 0.05 | -0.04,0.15 | 0.03 | -0.10,0.16 | 0.13 | -0.05,0.31 | **0.24** | **0.02,0.46** | **0.33** | **0.03,0.63** |
| Partly skilled/unskilled | **0.18** | **0.08,0.28** | **0.17** | **0.03,0.32** | **0.43** | **0.24,0.62** | **0.54** | **0.28,0.80** | **0.83** | **0.45,1.20** |
| **Educational Attainment** |  |  |  |  |  |  |  |  |  |  |
| University degree | Ref |  |  |  | Ref |  | Ref |  | Ref |  |
| GCE/School leaving cert | -0.01 | -0.11,0.09 | -0.05 | -0.18,0.09 | 0.02 | -0.18,0.22 | 0.14 | -0.10,0.38 | **0.35** | **0.03,0.67** |
| None | 0.06 | -0.07,0.18 | 0.13 | -0.04,0.30 | **0.28** | **0.04,0.53** | **0.62** | **0.32,0.91** | **1.08** | **0.67,1.49** |
| **Sex** |  |  |  |  |  |  |  |  |  |  |
| Men | Ref |  | Ref |  | Ref |  | Ref |  | Ref |  |
| Women | **0.09** | **0.02,0.15** | **0.15** | **0.06,0.24** | 0.06 | -0.07,0.18 | -0.11 | -0.27,0.06 | **-0.24** | **-0.46,-0.03** |

<sup>a</sup>All models adjusted for sex; coefficients in bold indicate 95% confidence intervals that do not include zero

**Figure 3.**
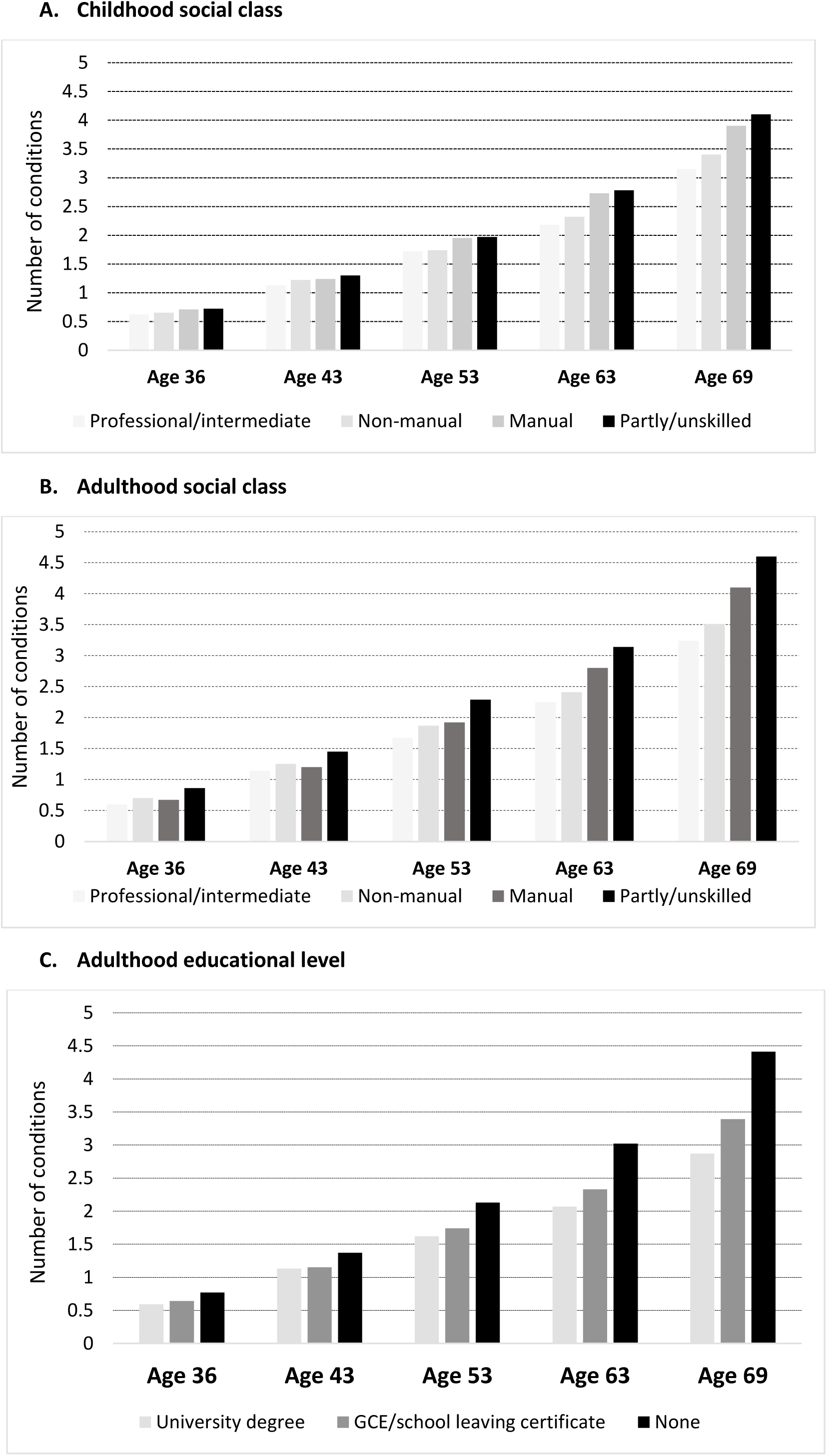
Mean number of morbidities at each age by socioeconomic indicators in 3,723 participants from the 1946 National Survey of Health and Development.

Overall, multimorbidity increased steadily over adulthood with an accelerated accumulation in older ages [from 0.08 (95% CI 0.07-0.09) conditions/year between ages 36-43 to 0.19 (0.18-0.20) conditions/year between ages 63-69 (linear spline mixed-effects models, Model 1, Supplemental Table 6, and Supplemental Figure 2)]. Further, we observed stark socioeconomic inequalities in multimorbidity trajectories which increased and widened progressively with age. Between ages 43 to 69, disadvantaged adulthood social class groups like manual and partly skilled/unskilled individuals experienced greater acceleration of multimorbidity trajectories (Model 3, Supplemental Table 6 and Figure 4A). For instance, compared to professional/intermediate individuals, manual workers experienced an excess of 0.18 conditions and partly skilled/unskilled individuals 0.31 conditions between ages 36-43 years, which increased to 0.30 conditions and 0.49 conditions respectively between 63-69 years (Model 3, Supplemental Table 6). The independent (of adult SEP) disparities in multimorbidity trajectories by childhood social class emerged at age 53 and widened over the remaining period of follow-up (Model 2, Supplemental Table 6 and Figure 4B). In this case, rather than a graded effect by childhood social class, professional and non-manual childhood social classes both followed a more favourable trajectory than manual and unskilled social classes.

**Figure 4.**
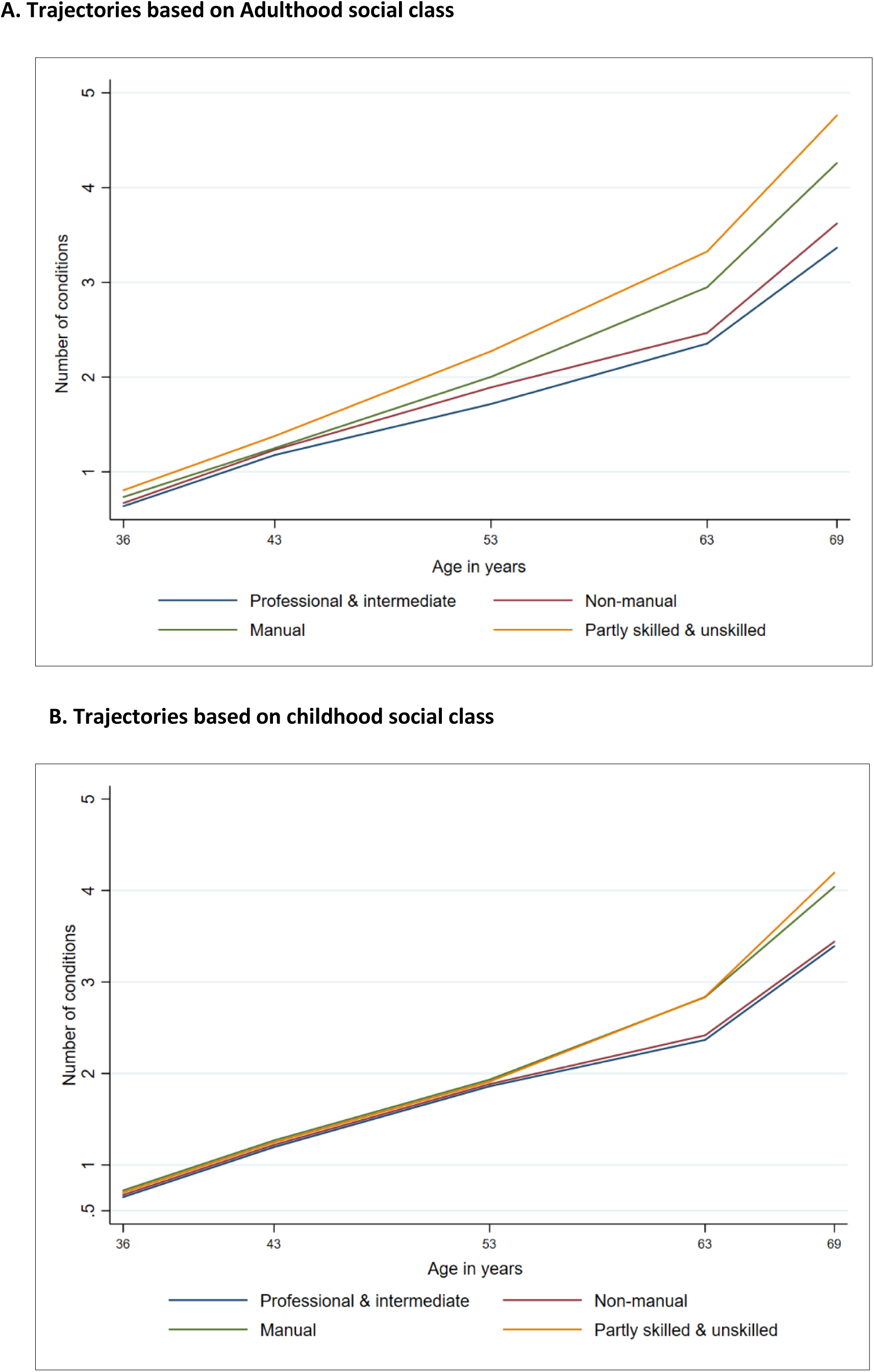
Socioeconomic inequalities in multimorbidity trajectories in 3,723 participants from the 1946 National Survey of Health and Development. Note: Figure 4 A is based on Model 3 in Table 3 and adjusted for sex, childhood social class and educational level. Figure 4 B is based on Model 2 in Table 3 and adjusted for sex, adulthood social class and educational level

## Discussion

Through following the same individuals in the longest running British birth cohort we report several novel findings. Firstly, we show that multimorbidity accumulation accelerates with age. Secondly, we find that childhood and adulthood socioeconomic position, and achieved education, each independently influence risk of multimorbidity burden at each age. Thirdly, we demonstrate that adverse socioeconomic status is associated with a more rapid acceleration of multimorbidity accumulation, providing strong evidence for widening inequalities with age. In sum, socioeconomically disadvantaged individuals have *both* earlier onset and more rapid accumulation of multimorbidity, with independent contributions from both childhood and adulthood socioeconomic position.

The few longitudinal studies that have examined associations between socioeconomic position and multimorbidity found that low income, low education, low occupational status and greater area deprivation were associated with increased risk for multimorbidity^10 16 21 29 30^. In addition to a shorter follow-up time, previous studies have focused on certain conditions^29^ (like cardiometabolic diseases) or analysed disease-specific trajectories^30^. Importantly, no previous study has examined socioeconomic patterning of multimorbidity trajectories over the lifecourse taking into account the independent effects of childhood and adulthood socioeconomic circumstances. Previous studies, for example using electronic health records in Finland, have also reported that inequalities increase with age.^10 21^However these may be misleading as ascertainment of chronic conditions, most of which are asymptomatic, in some cases for several years, may be biased towards either those with pre-existing morbidity, or worried well health care seekers. In capturing data on conditions from multiple sources, including self-report and standardised biomarker assessment on the whole cohort at multiple time points for the same individual, we overcome concerns over biased ascertainment.

In light of secular trends in health and life expectancy, and in widening childhood and adulthood socioeconomic inequalities, these findings have significant implications for more recent cohorts. These face much higher levels of multimorbidity as they age^2^ as a consequence of greater prevalence of earlier life obesity and mental ill-health and their sequelae, but also enhanced detection and lower thresholds for medication for conditions such as diabetes and hypertension and increased survival from life threatening conditions such as myocardial infarction and some cancers, all of which result in more years lived with multimorbidity.

### Strengths and limitations

This study utilises data on individuals followed-up from ages 36 to 69 enabling us to investigate the development of and increasing inequalities in multimorbidity trajectories using robust longitudinal modelling. Additionally, this study uses both periodical cross-sectional and longitudinal analyses to investigate socioeconomic patterning in multimorbidity. The NSHD was a nationally representative sample of all births in one week in the UK, with a >95% response rate. As with most longitudinal studies, the NSHD suffers from attrition. However, this study included 70% of the original birth cohort (N=5,362) and is reasonably representative of the British born population of the time.^31^ Nonetheless, the NSHD is composed of only members of white European heritage, and findings may not be generalizable to the non-White British population of the same generation. Some conditions like CHD, stroke and kidney disorders were defined by only self-report of a doctor’s diagnosis. However, this should not affect estimation of multimorbidity, as self-reporting of doctor’s diagnoses was not found to bias findings in other studies,^32 33^ and in NSHD >90% agreement for diagnosis of diabetes when comparing self-report with primary care diagnosis was shown.^34^ The population based nature of the study resulted in a comprehensive set of 18 conditions over time, however, studies based on electronic health records usually consist of a larger number of conditions.^10 21^ The inclusion of a limited range of common conditions and excluding more rare diseases in mid-adulthood is likely to have led to some underestimation of the extent of multimorbidity. The definitions and criteria for diagnoses of various diseases have changed over time making it difficult to study some conditions in greater detail, for example, all respiratory conditions had to be grouped together due to changing measurement methods and definitions over the last few decades. While there are several definitions of multimorbidity, we chose to use disease counts which is the most commonly used definition of multimorbidity enabling comparison to other studies and appropriate as multimorbidity is the outcome of interest (more complex definitions like weighted measures and indices are better suited to studying multimorbidity in relation to outcomes like mortality and healthcare usage).^35 36^

### Clinical and policy implications

The increase in multimorbidity in older ages is both a natural consequence of longer life expectancies and significant change in lifestyle seen in recent decades. By age 53, 52% of our study population experienced multimorbidity and we observe substantial inequalities in multimorbidity development. The largest increases in conditions over time were observed for those part of the cardiometabolic syndrome (obesity, hypertension, dyslipidemia and diabetes), and these conditions were more strongly interconnected over time as demonstrated by the network analysis. Primary care physicians are the main point of contact and ongoing care for patients with multimorbidity, with additional input from geriatricians with expertise in dealing with multimorbidity for older patients, and organ-specific clinical specialists for patients with specific conditions on a single disease basis. While there are limited clinical guidelines for addressing multimorbidity in the UK these as yet do not include any guidance on targeted care for disadvantaged individuals at higher risk for greater multimorbidity as clearly shown in this study{Farmer, 2016 #56}.

The independent and long-lasting shadow of childhood socio-economic position on both the experience and the rate of accumulation of multimorbidities in later life, highlight the need for interventions early in life to minimise inequalities in later life health. Importantly, the ongoing and additional inequities observed in relation to adulthood socioeconomic factors stress the need for intervention and prevention efforts starting in childhood, but also their continued provision through the lifecourse to reduce these age-widening inequities in health and their corresponding consequences for healthy ageing, years lived with disability and reduced life expectancy across all of society.

By 2035, the proportion of the UK population living with 4 or more conditions is expected to increase to 17% from 9.8% in 2015.^37^ Hypertension, chronic pain, obesity, depression and anxiety are expected to be the leading contributors to multimorbidity, indicating the heterogeneity of diseases likely to co-occur and the strong interlinking between mental and physical health conditions^3^. Given that conditions like obesity, depression and diabetes are socioeconomically patterned and more prevalent in younger generations, and combined with the projected increase in older age groups due to increasing life expectancies (those >75 years is expected to double to 13% over the next 20 years), the issue of multimorbidity, and its inequalities, will be a heavy burden that poses serious challenges for healthcare systems.

The synergistic clustering of diseases at both the population and individual level is one of the mechanisms by which health inequalities develop and perpetuate in sections of the population.^15 38 39^ Hence, any model developed to tackle multimorbidity and to reduce inequalities associated with socioeconomic circumstances should incorporate societal and individual-level factors. Additionally, public health policies that aim to reduce multimorbidity need to start earlier in the lifecourse, must be universal but proportional with increased targeting towards the more socioeconomically disadvantaged population.

### Final summary

In conclusion, this study shows marked and independent effects of childhood and adulthood socioeconomic circumstances on onset, rate of accumulation and burden of multimorbidity. These inequalities widen with time and the effects of socioeconomic disadvantage in childhood on multimorbidity emerges in late adulthood and early old age. Strikingly, the most socio-economically disadvantaged by each measure of SEP experienced an additional 0.39 conditions (childhood social class), 0.83 (adult social class) and 1.08 conditions (adult education) at age 69 years. Multimorbidity is perhaps inevitable in ageing societies with increasing life expectancies, but the inequalities as demonstrated by our study are stark. This not only calls for better access and delivery of healthcare for the more vulnerable, but also for population-based interventions in early life and through the lifecourse that reduce the impact of childhood inequalities if we are truly to address the societal impact of multimorbidity.

## Data Availability

Data are stored and held by the MRC Unit of Lifelong Health & Ageing, UCL. Requests for data can be made by contacting the unit directly.

## Ethics Approval

Written and/or informed consent was obtained from the study member at each stage of data collection.

### Funding

This study was funded by the following grants: This work was supported by the UK Medical Research Council which provides core funding for the MRC Unit For Lifelong Health and Ageing at UCL and the MRC National Survey of Health and Development (MC_UU_00019/1, MC_UU_00019/3). Amal Khanolkar was supported by grant from the Wellcome Trust (ISSF3/H17RCO/NG1) and MRC (MC_UU_00019/3). DB is supported by the Economic and Social Research Council (grant number ES/M001660/1) and The Academy of Medical Sciences/Wellcome Trust (“Springboard Health of the Public in 2040” award: HOP001/1025).

### Patient and Public Involvement

It was not appropriate or possible to involve patients or the public in the design, or conduct, or reporting, or dissemination plans of our research

### Competing interests declaration

NC is remunerated for her membership of a data safety and monitoring committee of a trial sponsored by AstraZeneca. The remaining authors have no competing interests to declare.

### Dissemination declaration

Results will be disseminated to study participants via the study newsletter and study website.

## Acknowledgements

We are grateful to the cohort members in the 1946 National Study of Health and Development who have provided data for research throughout their lives for their continuing support. We would like to thank the following clinicians who provided consultation and feedback: James Allinson, Gaby Captur, Sophie Eastwood, Samuel Searle, David Thompson and Alex Tsui.

We also thank members of the NSHD scientific and data collection teams.

## Author Contributions

PP and NC conceptualised the research aims with input from all authors and all authors contributed to the design of the study. AK prepared the dataset and conducted the analysis. VK conducted the network analysis. AK and PP drafted the paper with input from all authors. All authors discussed the findings, contributed to drafting and read and approved the final manuscript.

## Notes

### Author Declarations

Written and/or informed consent was obtained from the study member at each stage of data collection. The study obtained ethical approval from Greater Manchester Local Research Ethics Committee and the Scotland Research Ethics committee.

